# Incidence and Management of Early Breakthrough HSV Infection Among Allogeneic Hematopoietic Stem Cell Transplant Recipients

**DOI:** 10.64898/2026.08.02.26359522

**Authors:** Molly D. Fischer, Ria Mohan, Anna Wald, Amanda I. Phipps, Emily S. Ford, Ted Gooley, Frank Tverdek, Melinda A. Biernacki, Denise J. McCulloch, Michael J. Boeckh, Christine Johnston, Steven A. Pergam

## Abstract

**Background:** Reactivation of herpes simplex viruses (HSV) can occur in the early post-allogeneic hematopoietic cell transplant (aHCT) period despite antiviral prophylaxis. Few studies have assessed HSV infection in the modern era, in which acyclovir/valacyclovir is recommended for up to 1 year post aHCT. We evaluated the incidence and management of breakthrough HSV during the first 100 days post-aHCT over two decades.

**Methods:** Patients who received their first aHCT at Fred Hutchinson Cancer Center between 2002-2022 were reviewed for breakthrough HSV infection within the first 100 days on prophylaxis (acyclovir 800 mg or valacyclovir 500 mg twice daily). Cases were identified via culture, polymerase chain reaction, and/or direct fluorescent antibody testing; clinical records were reviewed for symptoms, outcomes, and prophylaxis/treatment regimens. Refractory/resistant (R/R) infections were defined according to consensus guidelines.

**Results:** We reviewed data from 4,357 aHCT recipients aged ≥18 years, among whom 3,749 (86%) were HSV seropositive and 23 developed breakthrough HSV infection (observed probability = 0.6%). Among those who had an infection, the median time from transplant to first positive test was 46 days (IQR: 24.0-69.5). Oral and genital mucosa were the most common sites of infection. In total, 11 of 23 (47.8%) patients with breakthrough HSV developed R/R infection.

**Conclusions:** Breakthrough HSV infections are rare in the first 100 days after aHCT among patients receiving antiviral prophylaxis. Refractory/resistant infections were uncommon but represented almost half of breakthrough cases. Our findings highlight the sustained effectiveness of universal prophylaxis in the early post-transplant period.

**Summary:** The incidence of breakthrough herpes simplex virus infections among adult allogeneic hematopoietic cell transplant recipients was low over a 20-year period at our center. Nonetheless, refractory/resistant infections represented almost half of breakthrough cases, highlighting the need for novel treatment strategies.

## Background

Prior to the introduction of standardized antiviral prophylaxis, herpes simplex virus (HSV) clinical disease occurred frequently among allogeneic hematopoietic cell transplant (aHCT) recipients.^1^ While HSV is primarily associated with recurrent self-limited oral or genital lesions in healthy persons, in immunosuppressed populations severe cases of non-healing oral or genital ulcers can occur, and disseminated infections can involve visceral manifestations such as pneumonitis^2^ or hepatitis.^3^ The use of acyclovir/valacyclovir prophylaxis for aHCT recipients has been officially recommended since 2000^4^ and has greatly reduced the risk of HSV disease. Long-term (i.e. at least 1 year) acyclovir/valacyclovir prophylaxis for varicella zoster virus (VZV) and HSV prevention has been used at Fred Hutchinson Cancer Center (FHCC) since 2002.^5,6^ Recent studies have evaluated the burden, outcomes, and associated treatment challenges of refractory/resistant HSV infection in the current era of standardized antiviral prophylaxis,^7,8^ but few have characterized the overall risk breakthrough HSV infection. The objective of this study was to evaluate the incidence and management of breakthrough HSV infection during the first 100 days post-aHCT.

## Methods

We retrospectively reviewed data from adult patients (aged ≥ 18 years) who underwent their first aHCT at FHCC, between January 2002 and December 2022. The aim was to identify cases of HSV within the first 100 days post-transplant, describe the clinical management of these cases, and identify potential risk factors for breakthrough HSV infection. We focused on the first 100 days because this is the most at-risk period for infections and other post-transplant complications such as acute graft-versus-host disease (aGVHD). Throughout the study period, standard VZV/HSV prophylaxis for aHCT recipients was twice daily 800 mg acyclovir or 500 mg valacyclovir for at least one-year post-transplant. Beginning in 2008, CMV seropositive umbilical cord blood (UCB) transplant recipients received high-dose (2g three times daily) valacyclovir for CMV prophylaxis;^9,10^ this continued until implementation of routine letermovir use in 2018.^11^

### Study Population

We reviewed center-specific databases for patients with positive HSV culture, polymerase chain reaction (PCR), or direct fluorescent antibody (DFA) results that occurred within 100 days of transplantation; samples were collected and sent for testing from various anatomic sites based on clinical discretion. Patients who had at least one positive test in this time period were defined as cases of HSV detection and selected for electronic health record (EHR) review. In a secondary analysis, we identified patients who had a second aHCT within 2 years of their index transplant (up until the end of 2022) to evaluate whether breakthrough HSV after first aHCT was associated with breakthrough HSV after subsequent transplants. To calculate incidence, the population at-risk of HSV infection was defined as patients seropositive for either HSV-1, HSV-2, or both by the University of Washington (UW) Western Blot (WB).^12^ The WB is the gold standard for type-specific HSV diagnosis, and was performed pretransplant for all patients.

### Data Collection

After demographic and transplant characteristics were extracted, EHR review for clinical aspects of presentation, including prophylaxis, symptoms, and treatment regimens was conducted. Test type (e.g. viral culture, PCR), HSV subtypes, anatomic site(s) and sample type for each positive test were characterized. The study was approved by the FHCC institutional review board.

### Definitions and Outcome Measures

#### HSV Detection, Breakthrough HSV Infection

HSV detection was defined as any positive HSV test result occurring in the first 100 days after transplant. Breakthrough HSV infection was defined as HSV detection in the setting of a lesion or other clinical signs of infection while the patient was receiving antiviral prophylaxis. Breakthrough HSV shedding was defined as any positive HSV test that did not present with visible lesions, was not treated, or was not deemed to be clinically significant by the care team. For cases where the symptom start date was not documented, we used the midpoint between the day of positive test and the last noted symptom-free day. For cases where symptom end date was not documented, we used the midpoint between the last day of symptoms and the first follow-up note that did not record symptoms.

#### Treatment and Antiviral Resistance

Refractory and/or resistant (R/R) HSV infections were defined according to consensus definitions.^13^ Refractory HSV lesions were defined as those that did not improve after at least one week of additional antiviral therapy or those where a new HSV-positive lesion developed after one week of treatment. Resistant HSV infection was defined as refractory HSV infection with phenotypic (performed via plaque reduction assay) or genotypic (sequencing) resistance to at least one antiviral drug documented in the EHR. In this study, patients who developed a breakthrough HSV lesion on prophylaxis that showed phenotypic or genotypic resistance were considered to have resistant HSV infection. If the treatment duration was not specified in the EHR, we assumed that the duration was two weeks.

### Statistical Analysis

Data extracted from the EHR were entered into Research Electronic Data Capture (REDCap).^14,15^ Descriptive statistics were used to summarize the incidence of HSV infection, infection characteristics, and patient outcomes. The chi square test was used to compare categorical demographic/transplant variables, and the Wilcoxon rank sum test was used for continuous variables. Due to the small number of breakthrough HSV cases relative to the total population size, we conducted a case-control study to investigate risk factors for breakthrough HSV infection. Controls were randomly selected from the population of HSV seropositive aHCT recipients via incidence density sampling (using the days since transplant) and individually-matched 3:1 to cases on age (within 5 years of the case) and calendar date of transplant (within 1 year of the case’s transplant). Potential risk factors selected *a priori* based on a review of the literature included sex, conditioning intensity, recipient CMV serostatus, and ≥ grade II aGVHD. Cell type (UCB vs. peripheral blood stem cells [PBSC] or bone marrow [BM]) were evaluated as potential risk factors after reviewing baseline transplant characteristics between patients with and without breakthrough HSV. We fit univariate conditional logistic regression models for each variable to evaluate the risk of breakthrough HSV infection. All analyses were conducted in RStudio version 2024.09.1+394.

## Results

### Patient Demographics

Between 2002 and 2022, a total of 4,357 patients received their first aHCT at FHCC. Among these patients, 3,749 were HSV seropositive (86%) and thus considered at risk of reactivation (**Table 1**). Among seropositive patients, 3,461 were HSV-1 seropositive (92.3%), 1,136 were HSV-2 seropositive (30.3%), and 848 (22.6%) were seropositive for both HSV-1 and HSV-2. The median age of seropositive patients was 53.8 (range: 18.5-80.9) and acute myeloid leukemia (AML) was the most common pre-transplant malignancy (40.0%). Half (12 of 23; 52.2%) of patients with breakthrough HSV had active aGVHD (≥ grade II) prior to HSV detection and most (17; 70.8%) developed aGVHD within 100 days of transplant (**Table 2**).

**Table 1:**
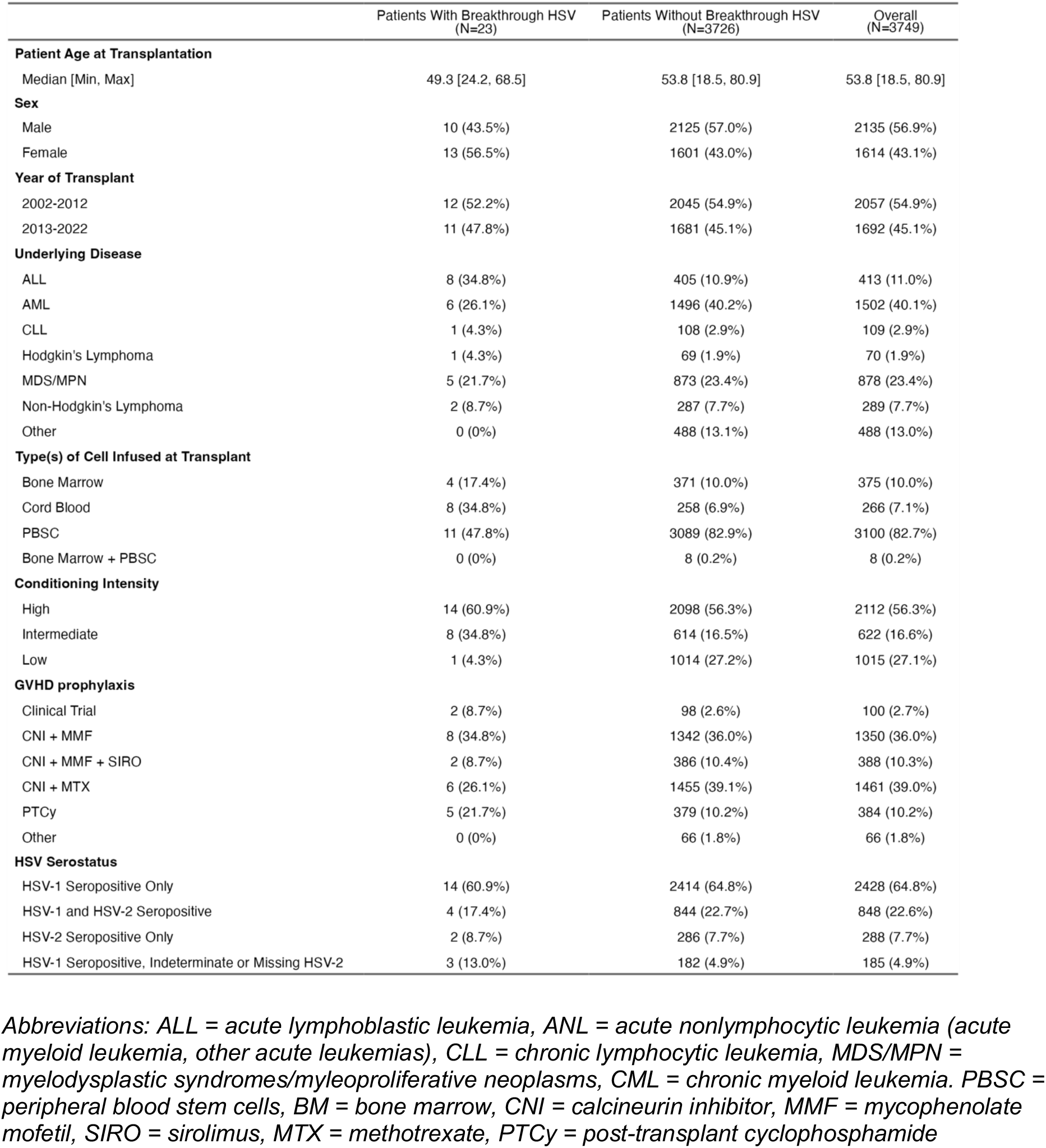
HSV Seropositive Patient Characteristics: With Breakthrough HSV Infection, Without Breakthrough HSV Infection, and Overall.

|  | Patients With Breakthrough HSV<br>(N=23) | Patients Without Breakthrough HSV<br>(N=3726) | Overall<br>(N=3749) |
| --- | --- | --- | --- |
| **Patient Age at Transplantation** |  |  |  |
| Median [Min, Max] | 49.3 [24.2, 68.5] | 53.8 [18.5, 80.9] | 53.8 [18.5, 80.9] |
| **Sex** |  |  |  |
| Male | 10 (43.5%) | 2125 (57.0%) | 2135 (56.9%) |
| Female | 13 (56.5%) | 1601 (43.0%) | 1614 (43.1%) |
| **Year of Transplant** |  |  |  |
| 2002-2012 | 12 (52.2%) | 2045 (54.9%) | 2057 (54.9%) |
| 2013-2022 | 11 (47.8%) | 1681 (45.1%) | 1692 (45.1%) |
| **Underlying Disease** |  |  |  |
| ALL | 8 (34.8%) | 405 (10.9%) | 413 (11.0%) |
| AML | 6 (26.1%) | 1496 (40.2%) | 1502 (40.1%) |
| CLL | 1 (4.3%) | 108 (2.9%) | 109 (2.9%) |
| Hodgkin's Lymphoma | 1 (4.3%) | 69 (1.9%) | 70 (1.9%) |
| MDS/MPN | 5 (21.7%) | 873 (23.4%) | 878 (23.4%) |
| Non-Hodgkin's Lymphoma | 2 (8.7%) | 287 (7.7%) | 289 (7.7%) |
| Other | 0 (0%) | 488 (13.1%) | 488 (13.0%) |
| **Type(s) of Cell Infused at Transplant** |  |  |  |
| Bone Marrow | 4 (17.4%) | 371 (10.0%) | 375 (10.0%) |
| Cord Blood | 8 (34.8%) | 258 (6.9%) | 266 (7.1%) |
| PBSC | 11 (47.8%) | 3089 (82.9%) | 3100 (82.7%) |
| Bone Marrow + PBSC | 0 (0%) | 8 (0.2%) | 8 (0.2%) |
| **Conditioning Intensity** |  |  |  |
| High | 14 (60.9%) | 2098 (56.3%) | 2112 (56.3%) |
| Intermediate | 8 (34.8%) | 614 (16.5%) | 622 (16.6%) |
| Low | 1 (4.3%) | 1014 (27.2%) | 1015 (27.1%) |
| **GVHD prophylaxis** |  |  |  |
| Clinical Trial | 2 (8.7%) | 98 (2.6%) | 100 (2.7%) |
| CNI + MMF | 8 (34.8%) | 1342 (36.0%) | 1350 (36.0%) |
| CNI + MMF + SIRO | 2 (8.7%) | 386 (10.4%) | 388 (10.3%) |
| CNI + MTX | 6 (26.1%) | 1455 (39.1%) | 1461 (39.0%) |
| PTCy | 5 (21.7%) | 379 (10.2%) | 384 (10.2%) |
| Other | 0 (0%) | 66 (1.8%) | 66 (1.8%) |
| **HSV Serostatus** |  |  |  |
| HSV-1 Seropositive Only | 14 (60.9%) | 2414 (64.8%) | 2428 (64.8%) |
| HSV-1 and HSV-2 Seropositive | 4 (17.4%) | 844 (22.7%) | 848 (22.6%) |
| HSV-2 Seropositive Only | 2 (8.7%) | 286 (7.7%) | 288 (7.7%) |
| HSV-1 Seropositive, Indeterminate or Missing HSV-2 | 3 (13.0%) | 182 (4.9%) | 185 (4.9%) |

*Abbreviations: ALL = acute lymphoblastic leukemia, ANL = acute nonlymphocytic leukemia (acute myeloid leukemia, other acute leukemias), CLL = chronic lymphocytic leukemia, MDS/MPN = myelodysplastic syndromes/myeloproliferative neoplasms, CML = chronic myeloid leukemia. PBSC = peripheral blood stem cells, BM = bone marrow, CNI = calcineurin inhibitor, MMF = mycophenolate mofetil, SIRO = sirolimus, MTX = methotrexate, PTCy = post-transplant cyclophosphamide*

**Table 2:** Outcomes of Patients with Breakthrough HSV Compared to Patients without HSV.

|  | Patients With Breakthrough HSV<br>(N=23) | Patients Without Breakthrough HSV<br>(N=3726) | Overall<br>(N=3749) |
| --- | --- | --- | --- |
| **Days From Transplant to Acute GVHD Onset** |  |  |  |
| Median [Min, Max] | 27.0 [7.00, 112] | 29.0 [0, 376] | 29.0 [0, 376] |
| Missing | 4 (17.4%) | 1041 (27.9%) | 1045 (27.9%) |
| **aGVHD Grade 2 or Higher within 100 Days** |  |  |  |
| No | 6 (26.1%) | 1271 (34.1%) | 1277 (34.1%) |
| Yes | 17 (73.9%) | 2455 (65.9%) | 2472 (65.9%) |
| **Death Within 1 Year of Transplant** |  |  |  |
| No | 14 (60.9%) | 2586 (69.4%) | 2600 (69.4%) |
| Yes | 9 (39.1%) | 1130 (30.3%) | 1139 (30.4%) |
| LTFU | 0 (0%) | 10 (0.3%) | 10 (0.3%) |
| **Relapsed Within 1 Year of Transplant** |  |  |  |
| No | 19 (82.6%) | 2949 (79.1%) | 2968 (79.2%) |
| Yes | 4 (17.4%) | 777 (20.9%) | 781 (20.8%) |

*Abbreviations: LTFU = lost to follow-up, aGVHD = acute graft-versus-host disease*

### Incidence of HSV and Risk Factors for Breakthrough Infection

A total of 1,620 HSV tests (excluding tests for viral resistance) were performed in the first 100 days post-transplant for this cohort, and almost half (741, 45.7%) were performed in the first 30 days. Among the 3,749 total HSV seropositive patients in this cohort, 824 (22.0%) were tested for HSV within the first 100 days (**Figure 1**). The number of tests performed and proportion of HSV seropositive patients tested per year are included in **Supplemental Figure 1**. Among these 824 patients, 31 tested positive (3.8%): 6 patients had HSV-2 detected, 16 had HSV-1 detected, and one had HSV without type specified detected. Among the 31 patients with HSV detection, 23 had breakthrough HSV infection and the remaining 8 had breakthrough shedding only. The cumulative incidence of type-specific breakthrough HSV infection was similar for HSV-1 and HSV-2; that is, 6 of 1,136 HSV-2 seropositive patients had breakthrough infection (0.5%) and 16 of 3,461 HSV-1 seropositive patients (0.5%) had breakthrough infection. The time to positive test and course of infection for patients with breakthrough HSV infection are shown in **Figure 2**. The median time from transplant to first positive HSV test among those with breakthrough HSV infection was 50 days (interquartile range [IQR] 36-80; range: 10-99 days),

**Figure 1:**
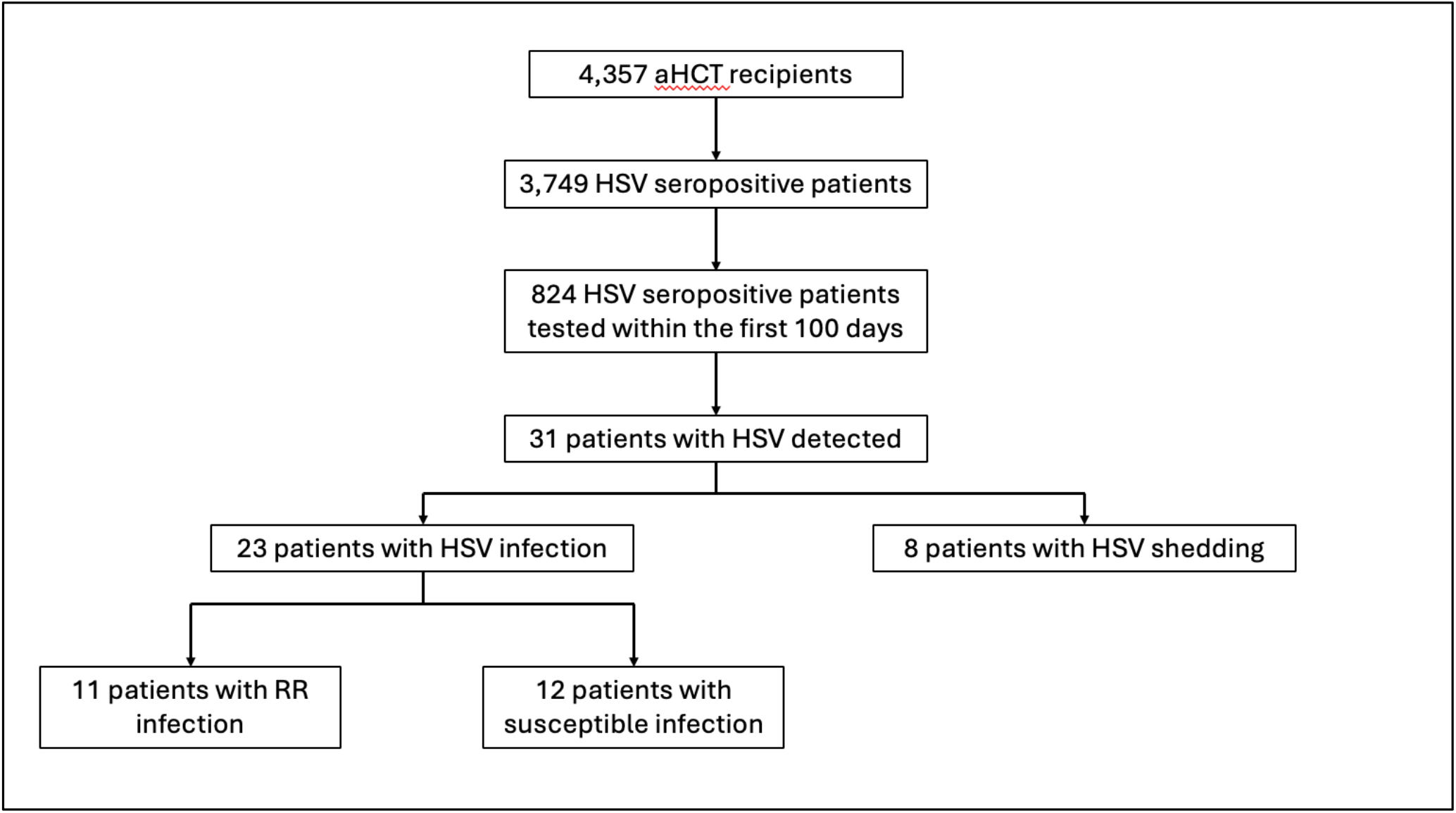
Patient flow diagram.

**Figure 2:**
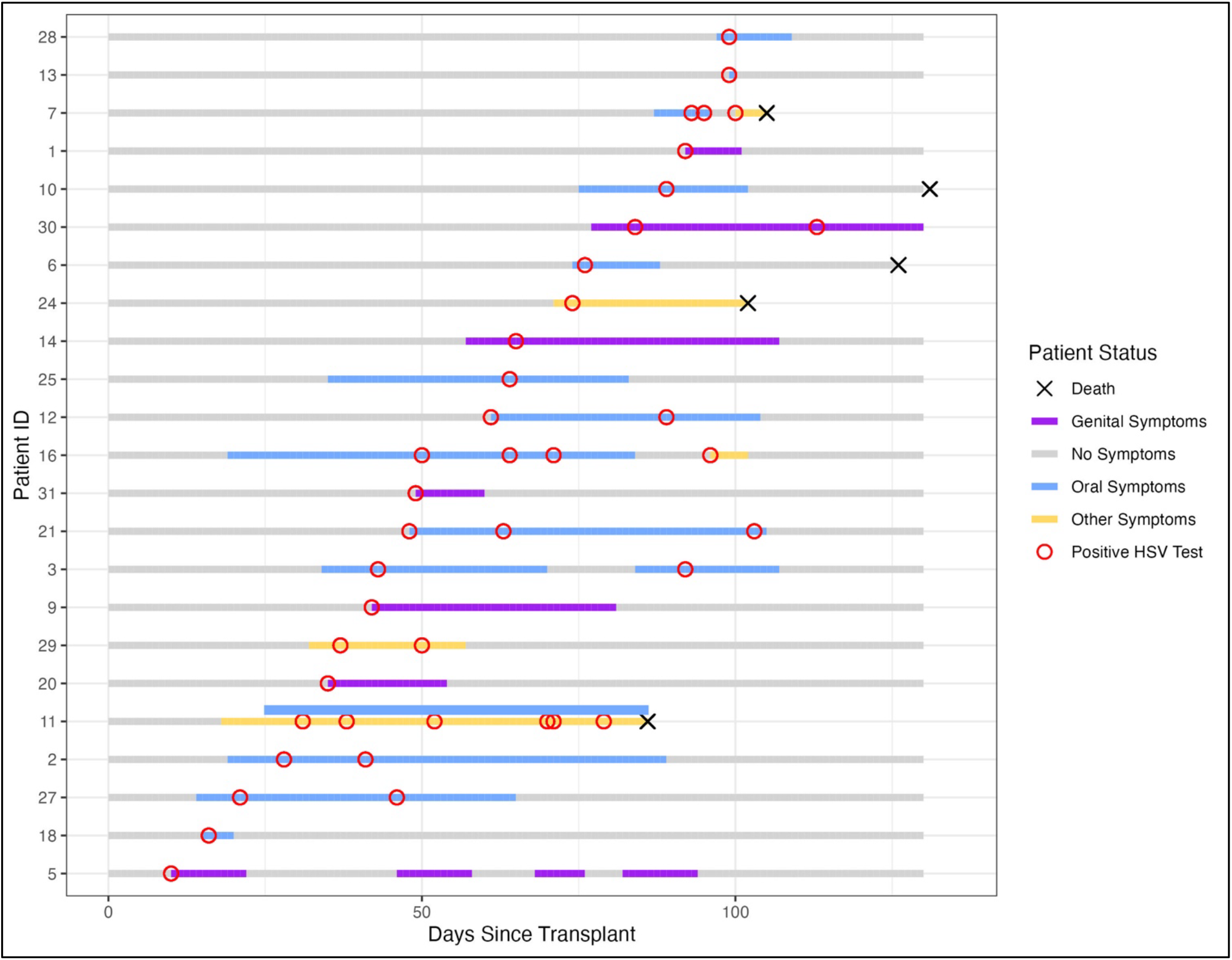
Time to first positive HSV test and clinical course: patients with breakthrough HSV infection (N = 23)* Clinical follow-up notes were unavailable for Patient 13 (oral lesion), and therefore symptom start and end dates could not be estimated and are not shown. Patient 11 experienced both oral and gastrointestinal symptoms leading to HSV detection, indicated by two symptom bars above. Type of HSV detected: All patients with genital HSV symptoms had HSV-2 detected except for Patient 31, who had genital HSV-1 infection. Patient 24 had nonspecific HSV type detected on CSF. All other patients had HSV-1 infection.

There were no statistically significant differences in pretransplant characteristics between patients with breakthrough HSV infection and seropositive patients without breakthrough, except for cell type (34.8% of patients with breakthrough infection received UCB compared to 6.9% of patients without; p < 0.001). The risk of breakthrough HSV infection among UCB recipients was 5.2 (1.5-17.4, p <0.01) times the risk of breakthrough HSV among PBSC or BM recipients. Sex, recipient CMV serostatus, or aGVHD were not significantly associated with breakthrough HSV infection (**Supplemental Table 1**).

### Sites of Detection and Symptoms

Among the 23 patients with breakthrough HSV infection, HSV was most frequently detected in the oral (n = 15 patients) and genital (n = 7) regions (**Figure 3)**. Additional sites of detection included the cerebrospinal fluid (CSF) (n = 1), stomach (n = 1), esophagus (n = 1), trachea (n = 1), lungs (n = 1), throat (n = 1), plasma/blood (n = 2) and eye (n = 1). Three patients had HSV detected in multiple locations. The time between transplant and symptom development for those with breakthrough infection varied widely (median 48 days [IQR 28.5- 74.5]).

**Figure 3:**
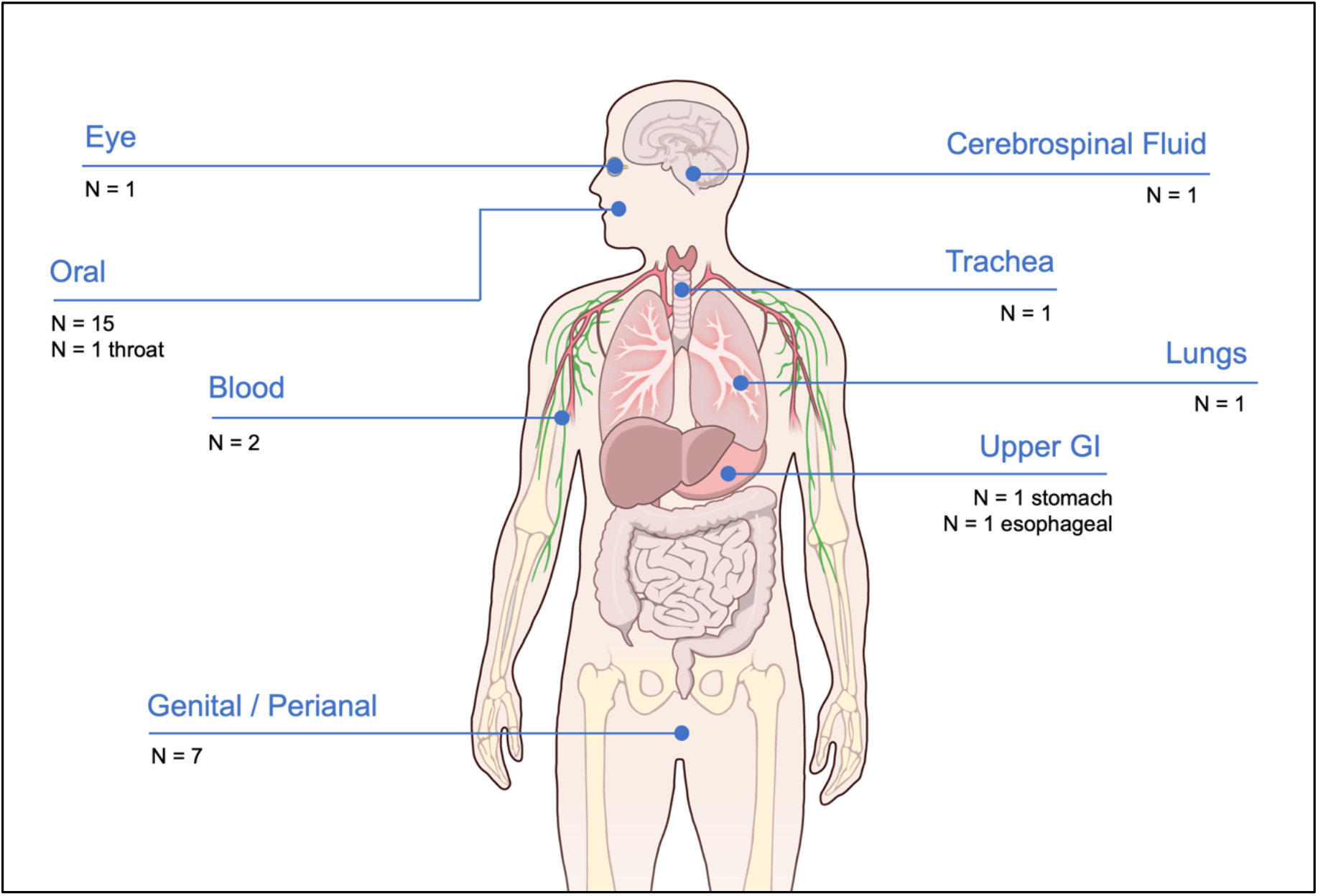
Anatomic Sites of HSV Detection Among Patients with Breakthrough Infection (N = 23) Illustration from *NIAID BioArt Source (bioart.niaid.nih.gov/bioart/519)*^32^ Counts reflect the number of patients with HSV detected at each site. Some patients had HSV detected at more than one anatomic site; total counts above therefore exceed the number (n = 23) of patients with breakthrough HSV infection. Patients with HSV shedding had nasal (n = 5), gastric (n =1), respiratory (n = 1), and bone marrow aspirate (n = 1) HSV detection (not shown).

Among the 8 patients with HSV shedding, HSV was detected on nasal wash (n = 5 patients), by bronchoalveolar lavage (n = 1), in bone marrow aspirate (n = 1), and in the stomach on gastroduodenoscopy (n = 1). This patient with HSV detection in the stomach was the only patient who was not HSV seropositive for the type-specific HSV detected. The patient was HSV-1 seropositive and HSV-2 seronegative but had culture-positive HSV-2 detected; the care team hypothesized that this was oral HSV-2 shedding and that the biopsy finding reflected procedural contamination.

### Outcomes

Breakthrough HSV infection was associated with a wide range of clinical presentations and outcomes (**Table 3**). Complications included refractory HSV esophagitis and gastritis (Patient 11), tracheitis (Patient 29), and ACV-resistant recurrent genital HSV-2 lesions (Patient 5). HSV infection was also considered a potential contributing cause of death for two patients (Patients 7 and 24). Patient 7 developed *Staphylococcus aureus* septicemia 10 days prior to their first positive HSV test from an oral ulcer; despite appropriate antibiotics, their condition worsened and they subsequently tested positive for HSV on BAL. Potential contributing causes of death listed included respiratory failure, renal failure, HSV, pneumonia and persistent bacteremia with severe GVHD; no autopsy was performed. Patient 24 developed severe HSV infection resulting in encephalitis; lumbar puncture revealed low levels of HSV (type nonspecific) in their cerebrospinal fluid. The patient had recently been transitioned to IV acyclovir due to difficulty with oral intake, which may have led to insufficient blood levels of ACV to inhibit HSV. In addition, the patient was thought to have posterior reversible encephalopathy syndrome, and experienced multiorgan failure (MOF); HSV PCR testing of serum was negative. They continued receiving acyclovir until death from presumed MOF and multifactorial (including HSV) encephalopathy. Overall, the incidence of relapse and all-cause mortality within 1 year of transplant was similar between patients with breakthrough HSV infection and seropositive patients without breakthrough infection (**Table 2**).

**Table 3:** Patients with Breakthrough Herpes Simplex Infections Within 100 Days of First aHCT.

| ID | Sex/<br>Age | Primary<br>Disease | Conditioning<br>Intensity;<br>Donor<br>Matching | HSV<br>Prophylaxis | HSV<br>Serostatus | Days<br>to 1<sup>st</sup> +<br>Test | Site<br>(HSV<br>Type) | Days to<br>GVHD<br>(Grade<br>≥ 2) | R/R<br>Infection | Treatment<br>(N Days) | Clinical Course |
| --- | --- | --- | --- | --- | --- | --- | --- | --- | --- | --- | --- |
| 1 | F/50's | MDS/<br>MPN | High;<br>MRD | Foscarnet<br>(Maintenance<br>Dose, 3g IV<br>daily) | HSV-1: (-)<br>HSV-2: (+) | 92 | Genital<br>(HSV-2) | 24 | No | VACV 500mg BID (5) | Lesion resolved on VACV |
| 2 | M/50's | ALL | High;<br>MURD | VACV 500mg<br>BID | HSV-1: (+)<br>HSV-2: (-) | 28 | Oral<br>(HSV-1) | N/A | Yes | 1. VACV 1g TID (7)<br>2. VACV 2g TID (10)<br>3. FOS 3.5g IV Q12<br>(14)<br>4. Pritelivir (25) (*trial*) | Refractory HSV that was<br>sensitive to ACV on phenotypic<br>assay. Grade III mucositis prior<br>to HSV detection. |
| 3 | F/40's | NHL | Intermediate;<br>DUCB | VACV 2g TID | HSV-1: (+)<br>HSV-2: (-) | 43 | Oral<br>(HSV-1) | N/A | No | FOS 5.4g IV Q12 (20) | Lesion resolved on foscarnet;<br>also had suspected herpetic<br>whitlow (lesion not tested, but<br>improved on foscarnet). Oral<br>recurrence ~1month later that<br>also resolved on foscarnet. |
| 5 | M/30's | MDS/<br>MPN | High;<br>MMURD | VACV 500mg<br>BID | HSV-1: (+)<br>HSV-2: (+) | 10 | Genital<br>(HSV-2) | 45 | Yes | 1. First lesion: ACV<br>(high dose, not<br>specified) (14)<br>2. Other lesions: FOS<br>(varying doses) (54) | Multiple breakthrough recurrent<br>genital lesions. First lesion was<br>ACV- and GCV-resistant;<br>recurrent lesions not tested. Last<br>recurrence required almost six<br>weeks of foscarnet. |
| 6 | M/50's | MDS/<br>MPN | Intermediate;<br>HAPLO | None | HSV-1: (+)<br>HSV-2: (-) | 76 | Oral<br>(HSV-1) | 22 | No | 1. VACV 1g TID (1)<br>2. GCV 450 mg IV<br>Q12 (10)<br>3. FOS 4.6 g =<br>191.67 mL IVPB Q24<br>Hours (14) | Oral lesions that appeared to<br>improve on foscarnet (GCV and<br>foscarnet primarily initiated for<br>concomitant CMV reactivation) |
| 7 | M/20's | ALL | High;<br>MMURD | ACV 500 mg IV<br>BID | HSV-1: (+)<br>HSV-2: (-) | 93 | Oral,<br>Respiratory<br>(HSV-1) | 23 | Yes | 1. ACV 500mg TID<br>(7)<br>2. FOS 4g (166.67<br>mL/hr on dialysis<br>days) (3) | Patient had *MRSA* infection that<br>did not resolve. ACV-resistant<br>oral HSV lesion, HSV on<br>subsequent BAL after intubation<br>for respiratory failure. HSV listed<br>as potential contributing cause of<br>respiratory failure and death. |
| 9 | F/30's | HD | Intermediate;<br>HAPLO | ACV 800mg<br>BID | HSV-1: (+)<br>HSV-2: (+) | 42 | Genital<br>(HSV-2) | N/A | Yes | 1. VACV 500mg TID<br>(22)<br>2. FOS 4.5g IV (21) | HSV-positive gluteal lesion and<br>rectal biopsy; ACV-resistant and did not fully heal on VACV. Resolved on foscarnet. |
| 10 | M/50's | CLL | High;<br>MURD | Foscarnet<br>(induction dose,<br>not further<br>specified) | HSV-1: (+)<br>HSV-2: (-) | 89 | Oral<br>(HSV-1) | 33 | No | VACV 500mg TID<br>(14) | Atypical-appearing HSV oral<br>ulcers that resolved on VACV. |
| 11 | F/40's | AML | High;<br>MRD | ACV 400mg<br>BID | HSV-1: (+)<br>HSV-2: (-) | 31 | Oral,<br>gastric,<br>esophageal<br>(HSV-1) | 38 | Yes | 1. ACV treatment<br>dose (not further<br>specified) (7)<br>2. FOS 2.4g IV daily<br>(54) | Stomatitis leading to discovery of<br>ACV-resistant infection.<br>Developed HSV esophagitis and<br>gastritis. Oral lesions and gastric<br>biopsy continued to be positive<br>until death. |
| 12 | M/40's | AML | Low;<br>MURD | ACV 800mg<br>BID | HSV-1: (+)<br>HSV-2: (-) | 61 | Oral<br>(HSV-1) | 25 | Yes | 1. VACV 500mg TID<br>(5)<br>2. VACV 1g BID (7)<br>3. VACV 500mg BID<br>(6)<br>4. VACV 1g TID (8)<br>3. FOS 2,688 mg BID<br>(11) | Refractory oral lesion that did not<br>improve on VACV, had additional<br>positive swab after ~1 month of<br>VACV. Rapid resolution on<br>foscarnet. |
| 13 | M/30's | MDS/<br>MPN | Intermediate;<br>HAPLO | VACV 500mg<br>BID | HSV-1: (+)<br>HSV-2: (-) | 99 | Oral<br>(HSV-1) | 20 | No | NA | Lip lesion; no follow-up notes<br>available |
| 14 | F/60's | AML | Intermediate;<br>DUCB | VACV 500mg<br>BID | HSV-1: (+)<br>HSV-2: (+) | 65 | Genital<br>(HSV-1) | 18 | Yes | 1. VACV 500mg TID<br>(12)<br>2. FOS 1.4g IV Q12<br>(23) | ACV-resistant genital lesion that<br>did not improve on VACV;<br>completely resolved on<br>foscarnet. |
| 16 | F/60's | NHL | Intermediate;<br>HAPLO | GCV 140mg IV<br>BID (for CMV) | HSV-1: (+)<br>HSV-2: (-) | 50 | Oral,<br>Throat<br>(HSV-1) | N/A | Yes | 1. VACV 1g TID (8)<br>2. FOS 60mg/kg/IV<br>BID (11) | ACV-resistant oral lesions along<br>with candida albicans; new HSV-1 culture-positive lesions<br>developed in throat while on<br>high-dose VACV. Lesions<br>resolved on foscarnet.<br>Had recently been switched to<br>GCV for CMV reactivation, but<br>oral symptoms began on VACV<br>(500mg BID). |
| 18 | F/30's | ALL | High;<br>DUCB | ACV 800mg<br>BID | HSV-1: (+)<br>HSV-2: (-) | 16 | Oral<br>(HSV-1) | 37 | No | ACV 800mg BID (no<br>additional treatment) | HSV-positive plaque in pharynx<br>that resolved on prophylactic<br>ACV without additional<br>treatment. |
| 20 | F/40's | ALL | High;<br>DUCB | VACV 500mg<br>BID | HSV-1: (+)<br>HSV-2: (+) | 35 | Genital<br>(HSV-2) | 44 | No | VACV 1g BID (14) | Labial cellulitis prior to lesion<br>development. Lesion resolved on<br>high-dose VACV. |
| 21 | M/50's | AML | High;<br>MURD | Foscarnet 3g<br>Q24 (for prior<br>HHV-6<br>encephalitis) | HSV-1: (+)<br>HSV-2: (-) | 48 | Oral<br>(HSV-1) | 20 | Yes | 1. ACV 400 mg IVPB<br>Q8 (19)<br>2. FOS 45 mg/kg Q12<br>(50) | ACV-resistant oral lesions. Initial improvement on ACV, lesions worsened and repeat culture was positive over 2 weeks later. Most but not all lesions resolved on foscarnet. Follow-up stopped at discharge. |
| 24 | F/40's | ALL | High;<br>DUCB | 800mg once<br>daily (due to<br>renal<br>insufficiency) | HSV-1: (+)<br>HSV-2: UN | 74 | CSF<br>(unspecifie<br>d HSV<br>type) | N/A | No | 800 IV Q12 Hours<br>(21) | Low levels of HSV in CSF by PCR. No serum HSV detected. Multifactorial encephalopathy, including HSV encephalitis, PRES, uremia, multiorgan failure leading to death. Suspected inadequate ACV intake due to challenges with taking oral medications. |
| 25 | F/40's | AML | High;<br>DUCB | VACV 2g TID<br>(following GCV<br>preemptive<br>therapy for<br>CMV) | HSV-1: (+)<br>HSV-2: (-) | 64 | Oral<br>(HSV-1) | 20 | No | 1. ACV 900 mg IV<br>Q12 (10)<br>2. GCV<br>(primarily for CMV)<br>(25) | Oral lesion that eventually resolved: initially treated with high-dose ACV, switched to GCV for CMV reactivation, resolved shortly after. Developed a new lesion (not tested) preceded by tingling and burning shortly after that improved while on GCV. |
| 27 | F/30's | ALL | High;<br>DUCB | ACV 400mg IV<br>Q12 | HSV-1: (+)<br>HSV-2: IND | 21 | Oral,<br>Plasma<br>(HSV-1) | 7 | Yes | 1. ACV 350mg IV Q8<br>(7)<br>2. FOS 6.5 grams IV<br>Q12 (6)<br>3. FOS 4.3 grams IV<br>Q12 (17)<br>3. VACV 500mg PO<br>TID (14) | Developed ACV-resistant lesions at prior oral mucositis sites; HSV viremia. |
| 28 | F/50's | ALL | Intermediate;<br>MURD | VACV 500mg<br>BID | HSV-1: (+)<br>HSV-2: (-) | 99 | Oral, Eye,<br>Blood<br>(HSV-1) | 27 | No | 1. VACV 500mg TID<br>(2)<br>2. VACV 1g TID (14) | Lesions on lip and buccal mucosa that resolved on VACV; HSV viremia. Bruising in eye prompted testing – HSV-1 was detected but determined not to be HSV infection due to lack of clinical signs of ocular HSV |
| 29 | M/60's | MDS/M<br>PN | Intermediate;<br>MURD | ACV 600mg IV<br>BID | HSV-1: (+)<br>HSV-2: (-) | 37 | Oral,<br>Tracheal<br>(HSV-1) | 54 | No | 1. ACV 7.5 mg/kg TID<br>IV (4)<br>2. FOS 3.45g Q24 (2)<br>3. FOS 1.78g Q24 (9) | Ongoing respiratory distress;<br>intubation leading to discovery of<br>oral and throat HSV lesions.<br>Diagnosed HSV tracheitis.<br>Empirically switched to foscarnet<br>over concerns of resistance (was<br>ACV-sensitive), lesions improved<br>on foscarnet. |
| 30 | M/50's | ALL | High;<br>DUCB | VACV 500mg<br>BID | HSV-1: (+)<br>HSV-2: (+) | 84 | Genital<br>(HSV-2) | 29 | Yes | 1. VACV 500mg TID<br>(28)<br>2. VACV 1g TID (22)<br>3. Pritelivir (41) (*trial*) | Rectal HSV-2 lesions initially<br>improved after ~2 weeks VACV,<br>then gradually worsened despite<br>increased VACV, leading to<br>discovery of ACV-resistant<br>infection. Patient received<br>pritelivir as part of clinical trial;<br>lesions resolved on pritelivir. |
| 31 | F/30's | AML | High;<br>MURD | ACV 800mg<br>BID | HSV-1: (+)<br>HSV-2: IND | 49 | Genital<br>(HSV-1) | NA | No | VACV 1g TID (12) | Genital HSV-1 lesion; resolved<br>on VACV |

*Patients 4, 8, 15, 17, 19, 22, 23, 26 had HSV shedding only and are not shown*
Abbreviations: AML = acute myeloid leukemia; ALL = acute lymphoblastic leukemia; MDS/MPN = myelodysplastic syndromes/myeloproliferative neoplasms; NHL = non-Hodkin's lymphoma, HD = Hodgkin's disease, CLL = chronic lymphocytic leukemia
MRD = Matched Related Donor; MURD = Matched Unrelated Donor; MMURD = Mismatched Unrelated Donor; DUCB = Double Umbilical Cord Blood, HAPLO = haploidentical
ACV = acyclovir; VACV = valacyclovir; FOS = foscarnet
IND = indeterminate; UN = uninterpretable

### Antiviral Prophylaxis Adherence and Absorption

While antiviral adherence was not measured directly, one patient (Patient 24) was suspected to have missed antiviral doses, though drug levels were not tested. One additional patient (Patient 6) had a gap in their antiviral coverage: after finishing treatment for CMV reactivation, the patient was not restarted on HSV prophylaxis until oral HSV lesions were identified approximately 2 weeks later.

### ACV Resistance, Clinical Management, and Outcomes

Patients treated for breakthrough ACV-susceptible infections received a median of 18 days of therapy (IQR: 13.5-24.5 days). Of the 11 patients who developed R/R breakthrough HSV infection, 9 had laboratory confirmed phenotypic/genotypic resistance and 2 had refractory HSV infection without documented resistance. The median length of treatment for R/R infections was significantly longer at 44 days (IQR: 37.5-62 days, p = 0.003). While V/ACV was used for primary HSV prophylaxis among this cohort, 4 patients developed breakthrough HSV infection while on antivirals primarily for prevention/treatment of other herpesviruses, such as CMV and HHV-6, including foscarnet (n = 3) and ganciclovir (GCV) (n = 1). Of these patients, 2 were receiving once-daily (maintenance) dosing of foscarnet and 1 was receiving twice-daily (treatment/induction) dosing at the time of positive HSV test. The patient on induction dosing developed R/R oral HSV-1. The patient who tested positive for HSV while on GCV had recently been switched from VACV, and symptoms began prior to initiating GCV.

### Secondary Analysis: HSV After Subsequent Transplant

A total of 85 HSV seropositive patients received a second transplant within 2 years of their first aHCT. Among the 31 patients with HSV detected after first aHCT, only one patient (Patient 20) had another transplant within 2 years. This patient had a history of oral and genital HSV and developed a genital HSV-2 lesion on Day 35 post-transplant that resolved on high-dose acyclovir after their first transplant. Following their second transplant approximately one year later, the patient developed a complicated course of refractory HSV infection beginning Day 21 post-transplant, including oral, throat, and esophageal HSV-1 lesions in addition to genital HSV-2 recurrence that persisted until death (unrelated to HSV) despite weeks of treatment.

## Discussion

This study provides an updated evaluation of the temporal trends and clinical manifestations of breakthrough invasive and mucocutaneous HSV infections in aHCT recipients while on acyclovir/valacyclovir prophylaxis. Our results suggest that the incidence of breakthrough HSV infection among aHCT recipients in the first 100 days post-transplant is very low. However, the high proportion of R/R infections among those with breakthrough HSV underscores the difficulty of managing HSV infections in this population.

While follow-up times, sampling strategies, and patient populations differ between published studies, our finding that the median time to post-transplant HSV detection (46 days) is consistent with those from studies among patients not on V/ACV^1,16^ and on antiviral prophylaxis,^17,18^ and with the high-risk for HSV reactivation in the early post-transplant period. In this study, the shorter median time to HSV shedding (18 days) compared to breakthrough infection (50 days) may reflect the greater number of tests performed in the early (i.e. first 30 days) post-transplant period, the time most associated with mucositis, or the ability of the virus to subclinically reactivate during the periods associated with more profound immunosuppression. The overall low incidence of breakthrough HSV in this study may also reflect the fact that FHCC doses acyclovir/valacyclovir prophylaxis at the upper end of the recommended range, and this may be higher than the doses used at other centers.^19^

When examining potential risk factors for breakthrough HSV infection, we found that UCB transplant recipients had a five-fold risk of breakthrough HSV relative to PBSC or BM recipients. This is consistent with prior research that suggests risk of reactivation of other herpesviruses such as CMV,^20,21^ human herpesvirus 6^22^ and VZV^23,24^ are higher in UCB recipients due to delayed immune recovery.^25,26^ Despite the high relative risk, the absolute risk of breakthrough HSV infection among seropositive UCB recipients on prophylaxis was low (3.0%).

The management of patients with R/R HSV remains a clinical challenge. Although R/R infections were overall uncommon, they represented almost half of breakthrough HSV cases in this cohort. R/R infections often caused substantial morbidity, prolonged hospitalization, and required weeks of high-dose antiviral therapy for patients to achieve symptom resolution. Several patients experienced regimen-related renal dysfunction. These complications require careful monitoring and underscore the importance of developing novel alternative antiviral therapies (e.g. helicase-primase inhibitors)^27^ for managing R/R HSV infections in immunocompromised populations.

The detection of HSV in other sites, such as the stomach and lungs, raises additional challenges for clinicians. While positive results could indicate true HSV infection, they may also reflect oral contamination from diagnostic procedures, including endoscopy/bronchoscopy that can introduce HSV into deeper anatomic sites. HSV PCR is highly sensitive, which may also lead to more frequent detection of low-level shedding. Because HSV can contribute to severe manifestations, such as esophagitis^28^ or encephalitis, it is crucial for clinicians to carefully consider the potential source(s) of reactivation as well as risks and benefits of antiviral therapy.

This study also has important limitations. Due to the retrospective nature of these data and overlapping clinical symptoms of HSV and conditions like mucositis, the start and end dates for HSV symptoms are estimates rather than exact dates. In addition, HSV may not have been in the differential diagnosis, and testing may not have been ordered in all cases. However, the low proportion of positive HSV tests also indicates that HSV testing may have been overutilized in this cohort. Protocols to limit testing could provide opportunities for diagnostic stewardship but require balance to assure identification of true breakthrough events. While we identified 2 cases of breakthrough infection that were potentially attributable to malabsorption, nonadherence to prophylaxis, or gaps in coverage as documented in the EHR, not all instances may have been identified and recorded, particularly with respect to adherence to outpatient antiviral regimens. Donor HSV serostatus is also not routinely assessed, but may be an important predictor of both breakthrough HSV infection incidence and severity of disease among HSV seropositive aHCT recipients.^29^

This study relied on symptom-driven sampling rather than routine testing, which precludes estimation of the frequency of subclinical HSV reactivation/shedding. While subclinical HSV reactivation is thought to occur at least once per month among immunocompetent populations, data on the frequency of asymptomatic HSV reactivation among aHCT recipients on antiviral prophylaxis are limited. In this study, the occasional detection of HSV without clinically-apparent manifestations suggests that subclinical shedding is occurring among aHCT recipients despite prophylaxis. A prospective study by Kakiuchi et al. of once weekly oral samples from HCT recipients found that 15% of patients had HSV-1 shedding detected at some point during the first 100 days.^30^ Subclinical shedding is also common among solid organ transplant recipients not on antiviral prophylaxis^31^, and likely occurs more frequently than among immunocompetent populations. The clinical significance of this shedding among aHCT recipients is unknown, but future studies incorporating regular frequent sampling intervals could address this knowledge gap.

## Conclusions

This large, single-center retrospective study underscores the effectiveness of acyclovir/valacyclovir prophylaxis for the prevention of early HSV infections after aHCT, while also highlighting the potential severity of breakthrough infections. Antiviral prophylaxis has dramatically reduced the incidence of HSV disease; however, challenges associated with treating breakthrough infection (particularly R/R infection) including regimen-related toxicity, support the need for novel therapies. Continued research into effective prophylactic and therapeutic strategies, alongside better diagnostic protocols, could further limit breakthrough HSV infection in aHCT populations.

## Data Availability

Additional data beyond what is offered within the full manuscript are not publicly available.

## Acknowledgements/Author Contributions

This work was supported by NCI NCI-P30CA0087-48. REDCap use was supported by the Institute of Translational Health Sciences at UW; NIH awards to the institute did not provide direct funding for this project. The authors thank Chris Davis, Agnes Ho, & Ryan Basom for their data support, and the HCT recipients who made this research possible. Additional data beyond what is offered within the full manuscript are not publicly available.

SAP, CJ conceptualized the study; SAP, CJ, MDF, TG, AW developed the methodology; MDF performed formal analysis; AW, ESF, FT, DM, MJB, CJ, SAP, MAB, RM, MDF conducted the investigation; RM, MDF wrote the original draft; all others reviewed and edited the final manuscript; RM, MF performed the data visualization; SAP, CJ provided supervision.

---

**Supplemental Table 1:** Odds Ratios and 95% Confidence Intervals for Risk Factors for Breakthrough HSV Infection.

| Variable | OR | 95% CI | P-value |
| --- | --- | --- | --- |
| Sex (male vs. female) | 0.45 | 0.17-1.16 | 0.10 |
| Conditioning intensity (low/intermediate intensity vs. high intensity) | 0.84 | 0.27-2.67 | 0.77 |
| Recipient CMV serostatus (seropositive vs. seronegative) | 1.13 | 0.42-3.04 | 0.80 |
| Prior CMV reactivation (yes vs. no) | 2.21 | 0.80-6.13 | 0.13 |
| aGVHD grade 2 or higher | 0.60 | 0.19-1.90 | 0.38 |
| Cell type (cord vs. PBSC/BM) | 5.16 | 1.53-17.42 | <0.01 |

*Because controls were selected using risk set sampling and the outcome of HSV infection was rare, the odds ratios from the model are interpreted as relative risks.*

**Supplemental Figure 1:**
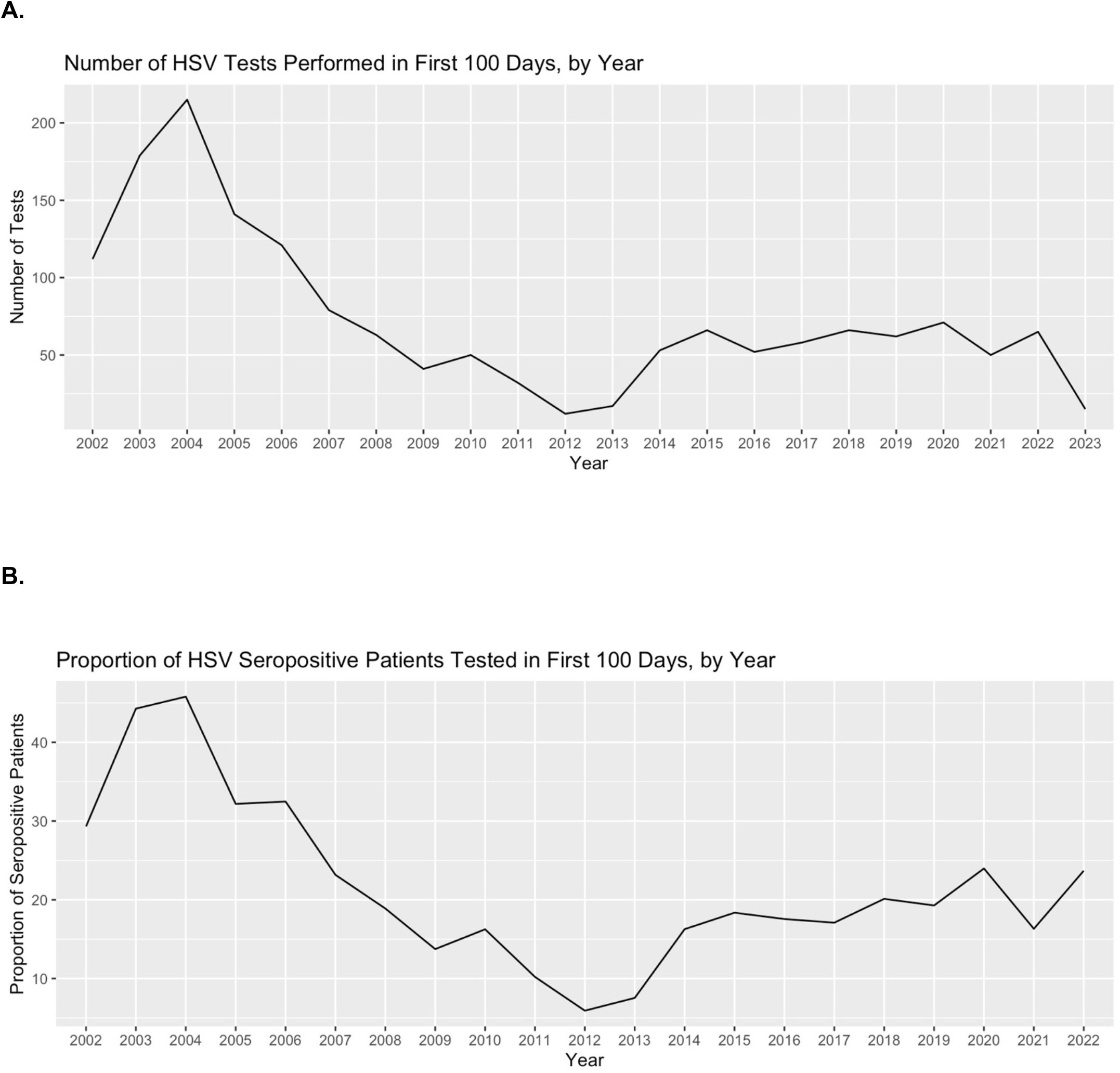
(A) Number of HSV Tests Performed and (B) Proportion of HSV Seropositive Patients Tested in the First 100 Days, by Year.

